# Optimizing scan efficiency of T1-weighted imaging for whole-brain intracranial vessel wall imaging

**DOI:** 10.1101/2025.06.23.25330079

**Authors:** Shraddha Pandey, Manuel Taso, Zhaoyang Fan, Konstanze V. Guggenberger, Alexia Tran, Scott B. Raymond, Shawn Lyo, Marisa Sanchez, Rob Sellers, Julien Savatovsky, Chengcheng Zhu, Quy Cao, Jae W Song, M. Dylan Tisdall

**Author notes:** Co-Corresponding Authors (J.W.S.), Perelman School of Medicine at the University of Pennsylvania, 3400 Spruce St, Philadelphia, PA 19104, United States., (M.D.T.), Perelman School of Medicine at the University of Pennsylvania, 3700 Hamilton Walk, Philadelphia, PA 19104, United States. Disclosures: MT and RS are employed by Siemens Medical Solutions, USA. Equal contributions (Jae W Song, M. Dylan Tisdall).

## Abstract

**Background:** Clinical intracranial vessel wall imaging (VWI) requires high spatial resolution leading to long scan times and artifacts.

**Purpose:** To accelerate standard-of-care (SOC) 3D T1-weighted variable-flip-angle turbo-spin-echo (VFA-TSE) sequence with parallel imaging (Generalized Autocalibrating Partially Parallel Acquisitions, GRAPPA) using compressed sensing (CS) or Controlled Aliasing in Parallel Imaging Results in Higher Acceleration (CAIPIRINHA, CAIPI) with either standard or large field-of-view (FOV) configurations to reduce scan time, artifacts and accommodate head sizes.

**Study Type:** Prospective study.

**Subjects:** Ten healthy volunteers.

**Field Strength/Sequence:** 3 Telsa, 20-channel head coil, T1-weighted VFA-TSE

**Assessment:** Accelerated sequences were compared to SOC GRAPPA (R=2), including standard FOV CAIPI (SFCAIPI, R=4), CS (SFCS7, R=7), and large FOV CS (LFCS7, R=7; LFCS10, R=10). Four neuroradiologists rated image quality (IQ) and signal-to-noise ratio (SNR) using a 4-point Likert scale. Scores of 3–4 were categorized as clinically interpretable. Lumen and wall diameters were measured.

**Statistical Analysis:** Descriptive statistics are reported. McNemar’s test compared proportions of IQ- and SNR-based clinically interpretable scans between relevant sequences of interest. Inter- and intra-rater reliabilities were calculated with Fleiss Kappa and weighted Cohen’s Kappa, respectively. Lumen and wall diameters of the CS- and CAIPI-accelerated sequences were compared to SOC using paired t-tests.

**Results:** SFCAIPI showed the lowest mean IQ and SNR scores. SFCS7 showed no significant difference in the proportion of IQ-based clinically interpretable scans compared to SFGRAPPA. When testing FOV, LFCS7 (35/40 scans; time of acquisition (TA)=3:45) showed a significantly higher proportion of IQ-based clinically interpretable scans compared to SFCS7 (27/40, p=0.03; TA=6:37). Upon increasing acceleration (R=10), there was no difference in the proportion of IQ-based clinically interpretable scans between LFCS7 and LFCS10 (36/40, p=0.65). Large FOV eliminated aliasing artifacts compared standard FOV (aliasing in 7 of 10 subjects). LFCS10 (TA=4:55) achieved a 50.6% reduction in TA relative to SFGRAPPA (TA=9:57).

**Conclusion:** Large FOV CS VWI sequence with 10x acceleration achieved a 50.6% reduction in scan time while delivering image quality comparable to SOC standard FOV GRAPPA.

## Introduction

Intracranial vessel wall MR imaging (VWI) is being increasingly performed to evaluate patients with intracranial vasculopathies and ischemic stroke (1). Optimal diagnostic imaging of intracranial VWI requires high spatial resolution (∼0.5 mm isotropic) with optimized tissue contrast and luminal blood flow suppression. However, these requirements typically require long acquisition times, increasing motion-related artifacts, and patient burden.

To address these challenges, previous studies explored acceleration techniques such as Generalized Autocalibrating Partially Parallel Acquisitions (GRAPPA), wave - Controlled Aliasing in Parallel Imaging (wave-CAIPI) (2), Controlled Aliasing in Parallel Imaging Results in Higher Acceleration (CAIPIRINHA, abbreviated as CAIPI) (3–4), and compressed sensing (CS) (5) to reduce scan times. In this study, we focused on acceleration methods supported for FDA-approved sequences available for our scanner, which include GRAPPA, CAIPI, and CS.

CAIPI and CS employ k-space sampling patterns that efficiently distribute aliasing artifacts caused by under-sampling, facilitating easier separation during image reconstruction. CAIPI uses a shifted sampling lattice that increases SNR in regions far from the receive coils (e.g., the center of the head near the Circle of Willis) when compared to similarly undersampled GRAPPA (3). CS approaches, on the other hand, use irregular sampling and leverage prior knowledge of the data by exploiting its sparsity in various transform domains, such as wavelet, finite differences, or Fourier domains combined with iterative regularized reconstructions (6, 7).

At our institution, a sagittally acquired 3D T1-weighted variable-flip angle Turbo-Spin-Echo (VFA-TSE) with parallel-imaging acceleration (e.g. GRAPPA) sequence is used for whole brain VWI protocols (8, 26). Non-selective excitation with a short hard pulse is used to reduce the echo time and maintain strong T1-weighting. As the human head in the left-to-right (LR) sagittal dimension is typically smaller than cranio-caudal or anterior-posterior (AP) dimensions, a sagittal orientation reduces acquisition time while achieving whole-brain coverage. To shorten acquisition times yet image key cerebrovascular anatomy, a standard field of view (FOV) measuring 136 mm in the LR dimension is planned, with the calvarium and ears covered by two slim saturation bands executed immediately before the excitation RF pulse to avoid strong aliasing. However, residual aliasing artifacts can be pronounced when imaging larger head sizes. Variability in head shapes and sizes among patients poses a challenge for clinical settings or when saturation is incomplete due to profile imperfection (9–11). Thus, a one-size-fits-all pulse sequence may not achieve the same image quality among all patients. To accommodate the range of patients’ head sizes/shapes, the FOV, among other parameters, may be adjusted by the scanning MR technologist during the exam (12). However, larger FOVs can adversely increase scan times and affect key imaging parameters such as spatial resolution and signal-to-noise ratio (SNR), inadvertently producing inconsistencies in image quality.

In this study, we test different acceleration techniques and large FOV, aiming to eliminate aliasing artifacts and accelerate scan times while maintaining diagnostic image quality. We hypothesize standard FOV CAIPI and CS acceleration techniques will not differ significantly from standard FOV GRAPPA VWI in quantitative and qualitative imaging assessments. We additionally anticipate that some of the acquisition time gained through advanced acceleration methods can, in turn, be traded for a larger imaging FOV, producing comparable image quality in less time than the SOC GRAPPA protocol.

## Materials and Methods

### Study Population

Ten healthy volunteers (3 females, 7 males, ages 43.8 ± 12.5 years) were prospectively recruited for this study. The study was approved by the local institutional review board. All subjects gave informed written consent for study participation. The data that support the findings of this study are available from the corresponding author upon reasonable request.

### Sequence Design and Magnetic Resonance Imaging

Imaging was performed on ten healthy subjects using a 3 Tesla MRI equipped with a vendor-supplied 20-channel head/neck coil (MAGNETOM Prisma Fit, Siemens Healthineers, Erlangen, Germany). The 20-channel coil was chosen as it is widely available across our health system and would facilitate implementation within our clinical workflow. Images were acquired in the sagittal plane with whole-brain coverage and 0.5 mm isotropic resolution. The readout direction was head-foot. We compared our clinical 3D VFA-TSE protocol using standard FOV (205×186×136 mm) GRAPPA R=2 (SFGRAPPA) with matched standard FOV protocols that differed only in the use of the acceleration methods: CAIPI R=4 (SFCAIPI) and CS R=7 (SFCS7). To address head size variability, we also tested the following large FOV (256×256×176mm) CS protocols: CS R=7 (LFCS7) and R=10 (LFCS10). All sequences had consistent voxel sizes (0.533mm isotropic), echo train lengths (ETL=52), and acquisition parameters (repetition time/echo time=900/16ms). Detailed scan parameters are listed in **Table 1**.

**Table 1.** Scan Parameters.

|  | SFGRAPPA | SFCAIPI | SFCS7 | LFCS7 | LFCS10 |
| --- | --- | --- | --- | --- | --- |
| Acquisition Time (mins:secs) | 9:57 | 5:51 | 3:45 | 6:37 | 4:55 |
| Reduced scan time (%) | NA | 41.2% | 62.3% | 33.5% | 50.6% |
| FOV (HF×LR×AP) mm | 205×136×186 | 205×136×186 | 205×136×186 | 256×176×256 | 256×176×256 |
| Voxel Size (isotropic) | 0.533 mm | 0.533 mm | 0.533 mm | 0.533 mm | 0.533 mm |
| ETL | 52 | 52 | 52 | 52 | 52 |
| TR/TE | 900/16 ms | 900/16 ms | 900/16 ms | 900/16 ms | 900/16 ms |
| Acquisition Orientation | Sagittal | Sagittal | Sagittal | Sagittal | Sagittal |
| Slices | 256 | 256 | 256 | 352 | 352 |

Abbreviations: Minutes (mins), Seconds (secs), Head Foot (HF), echo train length (ETL), Repetition time (TR), Echo time (TE), millimeter (mm), millisecond (ms), field of view (FOV), not applicable (NA), standard FOV GRAPPA R=2 (SFGRAPPA), standard FOV CAIPI R=4 (SFCAIPI), standard FOV CS R=7 (SFCS7), large FOV CS7 (LFCS7) and large FOV CS10 R=10 (LFCS10)

### Image Analysis

Four neuroradiologists with 4 to 9 years of experience interpreting intracranial VWI exams independently reviewed 50 randomized and anonymized scans from 10 healthy volunteers imaged with the 5 different acceleration techniques. For qualitative analyses, a 4-point Likert scale was used to evaluate the overall perceived SNR and Image Quality (IQ; 1 = poor, 2 = fair, 3 = good, 4 = excellent) (**Supplemental Table 1**). The basilar artery (BA), left and right internal carotid arteries (ICA, clinoid segment), and left and right middle cerebral arteries (MCA, M1 segment) were rated on each scan. Neuroradiologists were asked to provide an overall IQ rating for each artery along its entire segment. This was followed by asking the raters to provide IQ scores for the best and worst image slices along the length of the artery for a sensitivity analysis. To mitigate anchoring bias, each rater was asked to provide the overall IQ score first followed by scores for the best/worst segments. To assess intra-rater reliability, 4 neuroradiologists re-rated the overall image quality of 5 of the healthy volunteers (N=25 scans), blinded to the type of acceleration method after a ≥1-month wash-out period. Additionally, the presence of aliasing artifacts was scored visually when the ears or nose “wrapped around” the image in non-anatomic sites and projected at the LR or AP directions of the image, respectively. The anonymized and randomized pulse sequences were uploaded to PostDICOM (Herten, Netherlands), a cloud-based PACS viewer, for rating.

For quantitative analyses, two neuroradiologists (JWS, SL) blinded to the acceleration method, independently measured the lumen (inner-to-inner vessel wall) and wall (outer-to-outer) diameters of 3 of the vessels (BA, right or left ICA, and right or left MCA) **(Figure 1)**. Each diameter measurement was made in 2 axes (N= 600 lumen or wall diameter measurements for 3 vessels for 50 pulse sequences). Given the number of measurements and time-intensive nature of the task, unilateral MCA and ICA measurements were performed. To ensure measurement consistency between raters and account for the possibility of tortuous or tapering vessels (13), the coordinates for measurements of each vessel slice were pre-specified by plane and image numbers by one of the neuroradiologists and measurements replicated on the same slice by the second rater. If applicable, the neuroradiologist was also asked to note if the vessel inner or outer wall margins were not visible. For these cases, both neuroradiologists were asked to estimate a perceived diameter measurement based on image slices above/below and their anatomic knowledge, which allowed inclusion of all cases for quantitative analyses in a sensitivity analysis (14). The sensitivity analysis included 300 additional perceived measurements (N=150 lumen and 150 wall diameter measurements for the 3 vessels on the 50 scans). All measurements were made manually using 3D-Slicer (15).

**Figure 1.**
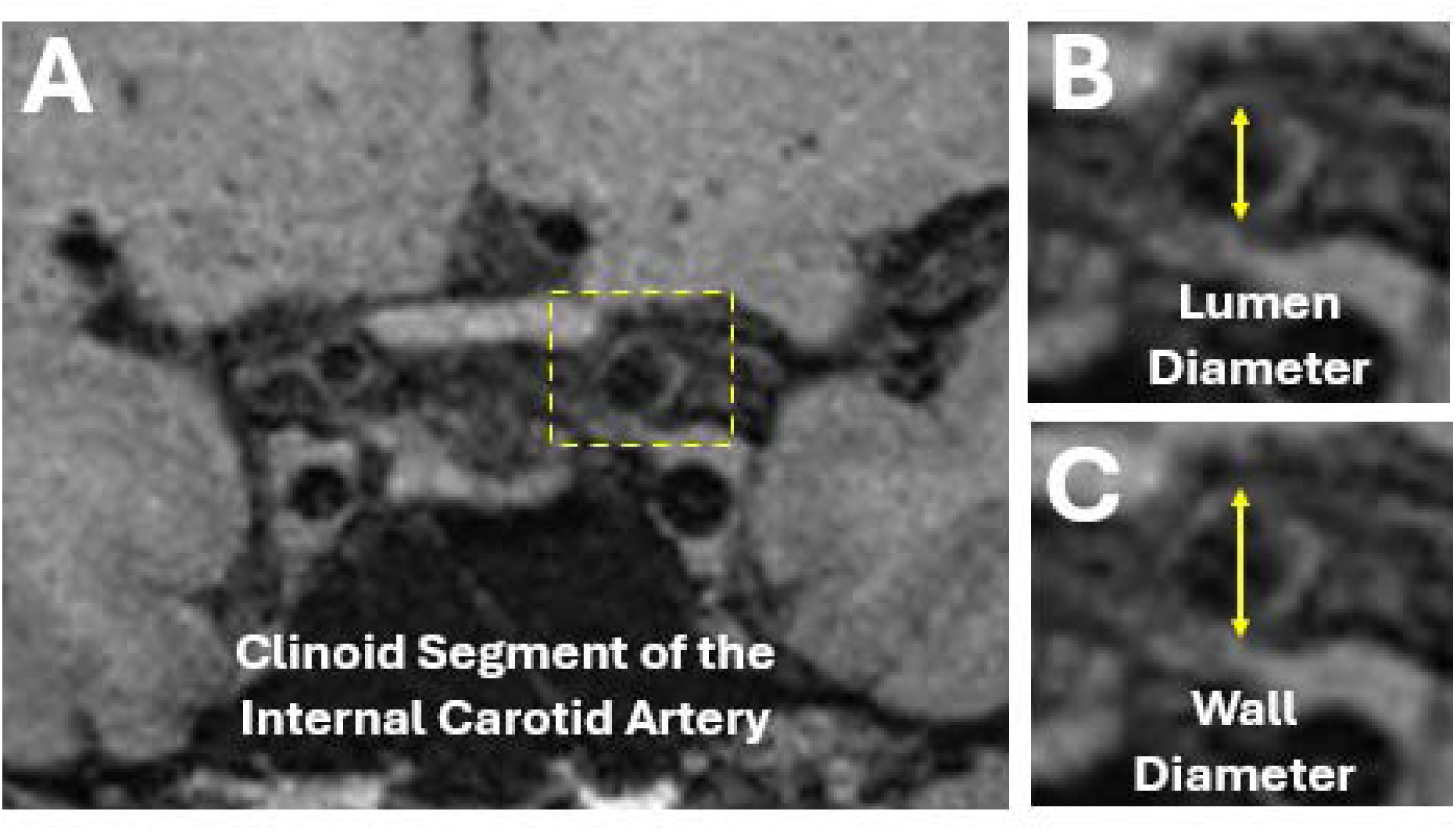
Examples of Left ICA Lumen and Wall Diameter Measurements Caption: (A) A coronal VWI of the intracranial internal carotid arteries are shown. The left clinoid ICA segment (dotted inset) is expanded in (B) and (C) to show how the lumen (inner wall-to-inner wall) and vessel wall (outer wall-to-outer wall) diameters were measured.

### Statistical Analysis

The qualitative scores among the four neuroradiologists are summarized as means and standard deviations. IQ and SNR scores were dichotomized for clinical interpretability, defined as scores ≥3 (interpretable) versus scores 1–2 (not interpretable), reflecting whether image noise adversely affected diagnostic confidence. McNemar’s test was performed on the dichotomized scores to compare the proportion of clinically interpretable scans between SFGRAPPA and each accelerated pulse sequence. Subgroup comparisons were also performed using McNemar’s test to assess differences in the proportion of clinically interpretable scans between standard and large FOV CS (SFCS7 vs LFCS7) and between different CS acceleration factors (LFCS7 vs LFCS10). The frequency of aliasing artifacts was also compared between standard and large FOV acquisitions.

Quantitative analyses were performed with the lumen and wall diameter measurements. The two measurements made for lumen and wall diameters were averaged for each vessel per rater. To account for variability in manual measurements, the mean of the averaged measurements from both neuroradiologists was calculated to obtain the final mean vessel lumen and wall diameters. When the diameter could not be measured by the neuroradiologist due to poor image conspicuity, the measurement was excluded from analysis. Sensitivity analyses with the perceived measurements were also performed. Paired t-tests were used to assess differences in the diameter measurements between SFGRAPPA and each accelerated sequence. Additionally, the number of lumen and wall diameter measurements per vessel that could not be measured by pulse sequence type was calculated.

Interrater reliability among the 4 neuroradiologists for image quality and SNR scores was assessed using the Fleiss Kappa Test. Inter-rater reliability was calculated using the weighted Cohen’s kappa test. Given the study’s exploratory nature, no formal correction for multiple comparisons was applied. Statistical significance was set as P <0.05. Analyses were computed using Matlab 2023b (Mathworks, Natick, MA).

## Results

### Qualitative Analysis: Image Quality of Whole Brain VWI

**Table 1** reports the acquisition times for the different acceleration methods. While maintaining the 0.533mm spatial resolution and standard FOVs, SFCAIPI and SFCS7 achieved 41.2% and 62.3% reductions in acquisition times compared to SFGRAPPA, respectively. Using large FOV parameters, LFCS7 and LFCS10 achieved 33.5% and 50.6% reductions in acquisition time, respectively.

**Table 2** summarizes IQ, SNR scores, and proportions of scans categorized as clinically interpretable. Among the standard FOV sequences, SFCAIPI had significantly lower image quality (1.48 ± 0.55) compared to SFGRAPPA, with a significantly lower proportion of IQ-based clinically interpretable scans (p=0.0005). In contrast, there was no significant difference in the proportion of IQ-based clinically interpretable scans between SFCS7 and SFGRAPPA (p=0.13). To assess the impact of FOV on CS-accelerated scans, SFCS7 and LFCS7 were compared, which showed SFCS7 had a significantly lower proportion of IQ-based clinically interpretable scans than LFCS7 (p=0.03). To evaluate the effect of acceleration, LFCS7 and LFCS10 were compared, for which there was no significant difference (p=0.65). Additionally, LFCS10 achieved the highest mean image quality score (2.95 ± 0.50) among all accelerated sequences while reducing scan time by 50.6% compared to SFGRAPPA.

**Table 2.**
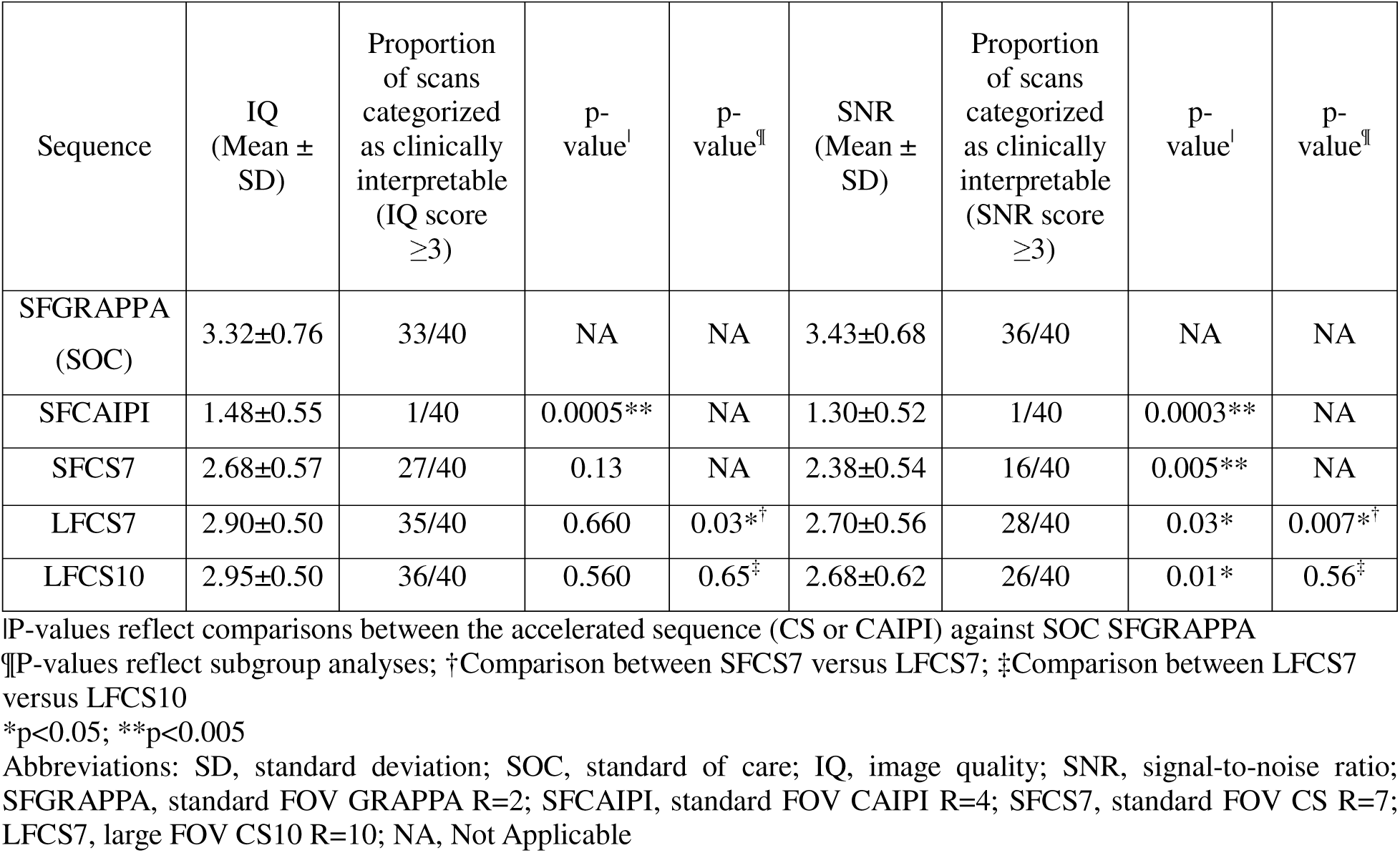
Image Quality (IQ) and Signal to Noise Ratio (SNR) Scores for the Whole Brain MR Image.

### Qualitative Analysis: Aliasing Artifact

**Figure 2** shows representative images of the basilar artery, middle cerebral artery, and internal carotid artery acquired using each sequence under standard or large FOV parameters. Aliasing artifacts were seen in 7 of 10 subjects using standard FOVs and were not observed in any subjects when imaged with large FOV parameters (**Figure 3**).

**Figure 2.**
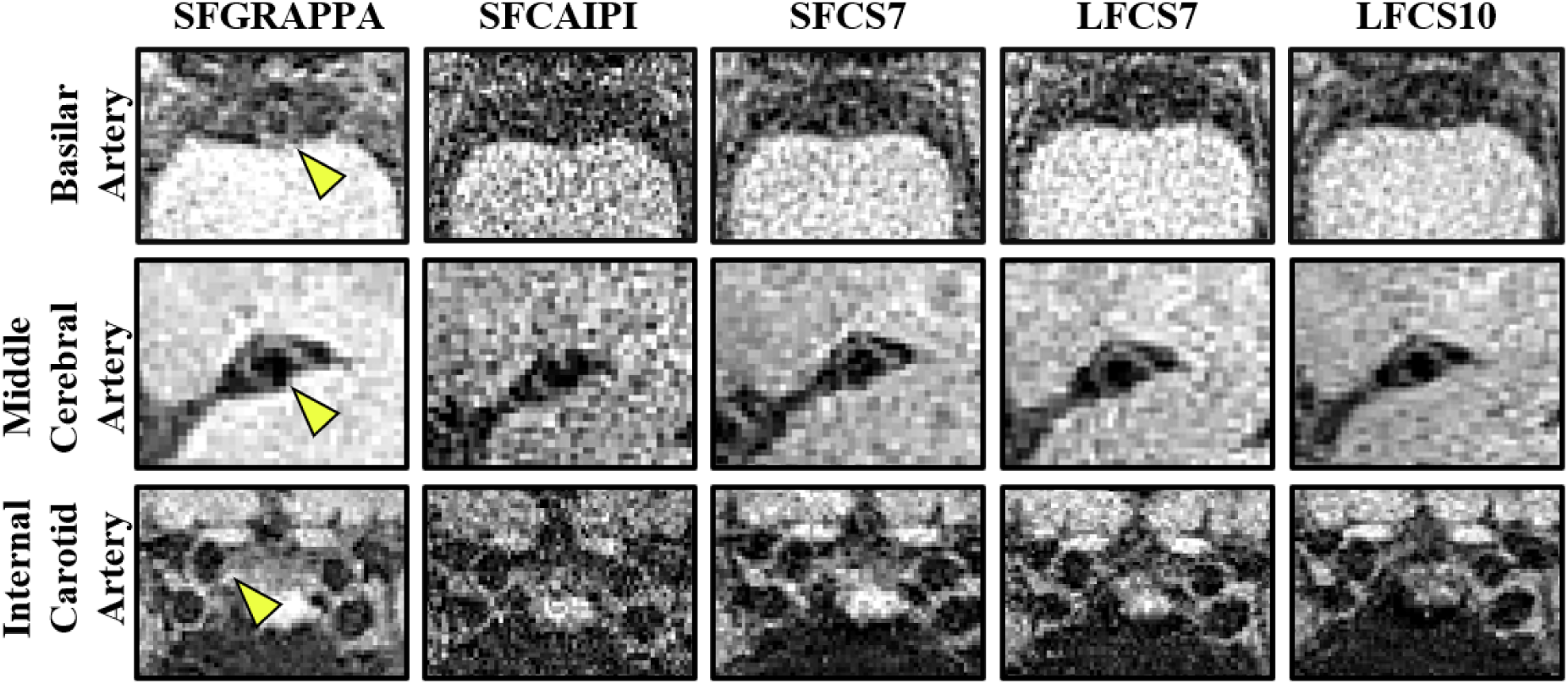
Representative Images of the Basilar, Middle Cerebral and Internal Carotid Arteries From the Five Accelerated Protocols Caption: Example images of the basilar artery (1^st^ row), middle cerebral artery (2^nd^ row), and internal carotid artery (3^rd^ row) at the level of the cavernous sinuses acquired from healthy subjects for the five protocols (left to right SFGRAPPA, SFCAIPI, SFCS7, LFCS7 and LFCS10) are shown. The yellow arrows indicate the vessel wall of the respective scored arteries. Abbreviations: SFGRAPPA, standard FOV GRAPPA R=2; SFCAIPI, standard FOV CAIPI R=4; SFCS7, standard FOV CS R=7; LFCS7, large FOV CS7; LFCS10, large FOV CS10 R=10

**Figure 3.**
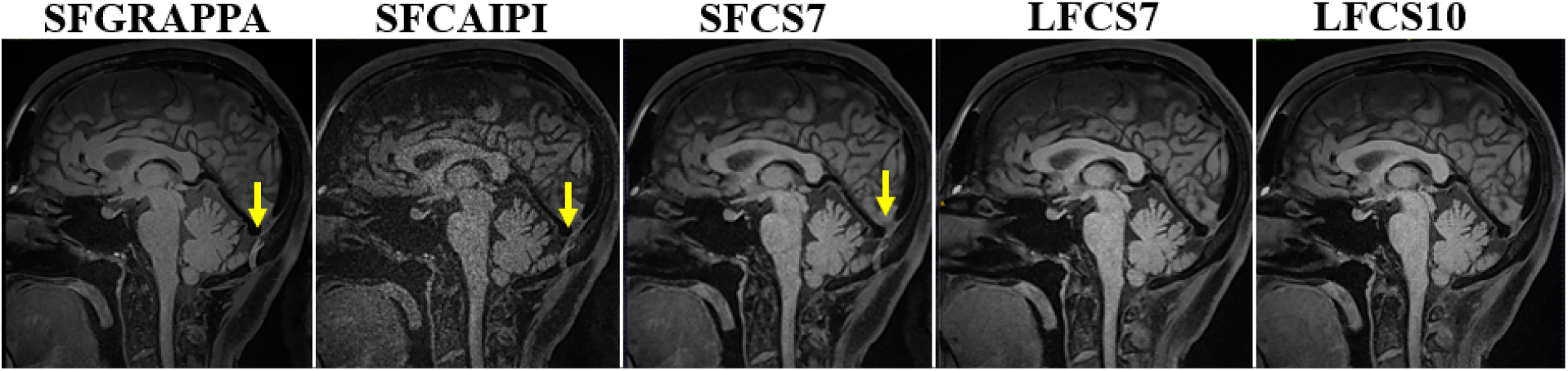
Presence of Aliasing Artifacts using Standard versus Large Field of View Image Parameters Caption: Example images of the acquired five pulse sequences for one subject. The yellow arrow shows the aliasing artifact on SFGRAPPA, SFCAIPI, and SFCS7. No aliasing artifact is observed on LFCS7 and LFCS10. Abbreviations: SFGRAPPA, standard FOV GRAPPA R=2; SFCAIPI, standard FOV CAIPI R=4; SFCS7, standar FOV CS R=7; LFCS7, large FOV CS7; LFCS10, large FOV CS10 R=10\

### Qualitative Analysis: Signal to Noise Ratio of Whole Brain VWI

All CS- and CAIPI-accelerated pulse sequences had significantly lower mean SNR scores compared to SFGRAPPA (Table 2). Subgroup analyses of the CS sequences comparing FOVs showed that the SNR score of SFCS7 (2.38 ± 0.54) was lower than that of LFCS7 (2.70 ± 0.56). Compared to LFCS7, SFCS7 also showed a significant difference in the proportion of SNR-based clinically interpretable scans (p=0.007). However, when comparing acceleration factors (R=7 vs. R=10), there was no significant difference in either SNR scores or the proportion of clinically interpretable scans between LFCS7 (2.70 ± 0.56) and LFCS10 (2.68 ± 0.62; p=0.56), indicating that increasing the CS acceleration factor did not further degrade SNR.

### Reliability Tests for Whole Brain VWI scores

For IQ of the whole brain, the interrater kappa value was 0.36 (95% CI 0.30-0.43) and intra-rater kappa value was 0.72 (95% CI, 0.63-0.81). For SNR of the whole brain, the interrater kappa value was 0.30 (95% CI 0.23-037) and intra-rater kappa was 0.73 (95% CI 0.63-0.81). These results indicate fair inter-rater and moderate intra-rater agreement for overall IQ and SNR, respectively.

### Qualitative Analysis: Image Quality Score by Vessel Segment

**Table 3** shows the mean IQ scores for each vessel segment from the four neuroradiologists. The BA consistently showed the lowest Overall Image Quality Score (OIQS) across all sequences compared to the MCAs and ICAs. For the BA, MCA, and ICA vessel segments, LFCS10 consistently scored the highest OIQS, and standard FOV CAIPI (R=4) scored the lowest. For the BA_OIQS_, all accelerated sequences (range: 1.13 ± 0.35 (SFCAIPI) to 2.43 ± 0.73 (LFCS10)) were statistically different compared to SFGRAPPA (2.77 ± 0.97). For the left and right MCA and ICA segments, all accelerated methods also showed statistically different and lower OIQS compared to SFGRAPPA. Thus, none of the accelerated pulse sequences showed comparable OIQS for the evaluated vessel segments compared to SFGRAPPA.

**Table 3.** Qualitative Overall Image Quality Scores for the Basilar, Middle Cerebral, and Internal Carotid Arteries.

|  | SOC | Accelerated VWI Sequences |  |  |  |
| --- | --- | --- | --- | --- | --- |
| Vessel | SFGRAPPA | SFCAIPI | SFCS7 | LFCS7 | LFCS10 |
| **Basilar Artery** |  |  |  |  |  |
| Overall | 2.77 ± 0.97 | 1.13 ± 0.35** | 1.97 ± 0.61** | 2.13 ± 0.73* | 2.43 ± 0.73 |
| Best | 3.08 ± 0.76 | 1.45 ± 0.50** | 2.40 ± 0.63** | 2.60 ± 0.71 | 2.58 ± 0.71* |
| Worst | 1.88 ± 1.04 | 1.03 ± 0.16** | 1.28 ± 0.51** | 1.43 ± 0.55** | 1.35 ± 0.62** |
| **Middle Cerebral Artery** |  |  |  |  |  |
| Left – Overall | 3.47 ± 0.73 | 1.50 ± 0.63** | 2.40 ± 0.67** | 2.70 ± 0.75 | 2.73 ± 0.64 |
| Left – Best | 3.43 ± 0.71 | 1.90 ± 0.74** | 2.68 ± 0.69 | 2.95 ± 0.6 | 3.03 ± 0.62 |
| Left – Worst | 2.63 ± 0.95 | 1.18 ± 0.45** | 1.78 ± 0.70** | 2.00 ± 0.82** | 1.98 ± 0.77** |
| Right – Overall | 3.50 ± 0.73 | 1.50 ± 0.63** | 2.37 ± 0.67** | 2.77 ± 0.68 | 2.80 ± 0.61 |
| Right – Best | 3.38 ± 0.74 | 1.90 ± 0.74** | 2.68 ± 0.62 | 2.93 ± 0.66 | 2.98 ± 0.53 |
| Right – Worst | 2.65 ± 1.00 | 1.18 ± 0.45** | 1.73 ± 0.68** | 2.10 ± 0.81* | 1.90 ± 0.81** |
| **Internal Carotid Artery (clinoid)** |  |  |  |  |  |
| Left – Overall | 3.33 ± 0.71 | 1.67 ± 0.61** | 2.63 ± 0.61 | 2.93 ± 0.69 | 2.97 ± 0.67 |
| Left – Best | 3.53 ± 0.64 | 1.98 ± 0.66** | 3.03 ± 0.58 | 3.23 ± 0.62 | 3.23 ± 0.58 |
| Left – Worst | 2.88 ± 0.88 | 1.28 ± 0.51** | 2.05 ± 0.55** | 2.30 ± 0.88* | 2.33 ± 0.86** |
| Right – Overall | 3.37 ± 0.72 | 1.63 ± 0.61** | 2.73 ± 0.58 | 2.97 ± 0.72 | 2.90 ± 0.66 |
| Right – Best | 3.55 ± 0.64 | 1.93 ± 0.69** | 3.03 ± 0.58 | 3.18 ± 0.64 | 3.15 ± 0.58 |
| Right – Worst | 2.93 ± 0.89 | 1.28 ± 0.51** | 2.13 ± 0.65** | 2.30 ± 0.82** | 2.33 ± 0.83** |

Caption: Values represent means and standard deviations. Comparisons are made between SFGRAPPA as the reference pulse sequence versus accelerated pulse sequences (SFCAIPI, SFCS7, LFCS7, and LFCS10). \*p<0.05; \*\*p<0.005
Abbreviations: SD, standard deviation; SOC, standard of care; SFGRAPPA, standard FOV GRAPPA R=2; SFCAIPI, standard FOV CAIPI R=4; SFCS7, standard FOV CS R=7; LFCS7, large FOV CS7; LFCS10, large FOV CS10 R=10

Sensitivity analyses were performed using the best and worst image quality scores. For the BA, MCA, and ICA, the SFCAIPI and SFCS7 sequences differed more frequently from SFGRAPPA compared to LFCS7 and LFCS10 sequences.

### Quantitative Analysis: Vessel Wall and Lumen Diameter

Among all pulse sequences, SFCAIPI was the least optimal for measuring vessel wall and lumen diameters due to poor conspicuity of vessel margins. Diameter measurements were not possible in 26.7% of evaluated vessels with SFCAIPI (16 missing measurements among the BA, MCA and ICA). The BA accounted for half of these (8 missing measurements), followed by the MCA (4) and ICA (4). In contrast, vessel margins were generally well visualized with SFGRAPPA, SFCS7, and LFCS7, with missing measurements in only 3.3% of evaluated vessels for each sequence. For LFCS10 and LFCS7, the ICA wall or lumen diameters were not visualized in 3.3% of vessels.

Using the diameters measured by the neuroradiologists, the MCA lumen and wall diameters on LFCS7 were larger and statistically different compared to the respective measurements on SFGRAPPA (**Table 4**). A sensitivity analysis using all measurements (actual measured and perceived) for lumen or wall diameters (**Supplemental Table 2**) showed additional significant differences compared to SFGRAPPA for the following outcomes: BA wall diameter on SFCAIPI, BA lumen diameter on SFCS7, and MCA lumen and wall diameters on LFCS7.

**Table 4.** Quantitative Values of Measured Lumen and Wall Diameters per Vessel Segment.

| **Vessel Lumen and Wall Diameter Measurements** | **SFGRAPPA (SOC)** | **SFCAIPI** | **SFCS7** | **LFCS7** | **LFCS10** |
| --- | --- | --- | --- | --- | --- |
| **BA- Lumen** | 2.98 ± 0.49 | 2.81 ± 0.37 | 3.14 ± 0.30 | 2.99 ± 0.27 | 3.01 ± 0.23 |
| **BA- Wall** | 4.86 ± 0.31 | 4.36 ± 0.76 | 4.89 ± 0.63 | 4.62 ± 0.44 | 4.94 ± 0.34 |
| **MCA- Lumen** | 2.58 ± 0.35 | 2.79 ± 0.49 | 2.57 ± 0.18 | 2.85 ± 0.34* | 2.60 ± 0.42 |
| **MCA- Wall** | 4.03 ± 0.49 | 4.25 ± 0.38 | 4.06 ± 0.34 | 4.35 ± 0.45* | 4.03 ± 0.62 |
| **ICA- Lumen** | 3.44 ± 0.79 | 3.60 ± 0.43 | 3.71 ± 0.40 | 3.59 ± 0.59 | 3.52 ± 0.69 |
| **ICA- Wall** | 5.29 ± 0.90 | 5.27 ± 0.62 | 5.49 ± 0.57 | 5.63 ± 0.59 | 5.40 ± 0.79 |

\*p<0.05; Values represent means ± standard deviations.
Abbreviations: SOC, standard of care; SFGRAPPA, standard FOV GRAPPA R=2; SFCAIPI, standard FOV CAIPI R=4; SFCS7, standard FOV CS R=7; LFCS7, large FOV CS7; LFCS10, large FOV CS10 R=10

## Discussion

We evaluated four CAIPI- and CS-based acceleration techniques applied to a 3D T1-weighted VFA-TSE sequence for intracranial VWI and compared them to our SOC GRAPPA-accelerated protocol. SFCAIPI performed worst with significantly lower observer-rated mean IQ and SNR scores and with nearly 25% of intracranial vessel segments deemed unmeasurable due to poor conspicuity. In contrast, CS-accelerated sequences were more frequently rated as clinically interpretable. To address head size variability, we compared standard versus large FOV CS-accelerated sequences. LFCS7 showed a significantly higher proportion of IQ-based clinically interpretable scans than SFCS7 (p=0.03) at longer scan times. To further reduce scan time, R=10 (LFCS10) was tested, which showed no significant difference in the proportion of IQ-based clinical interpretability scans compared to LFCS7 (p=0.65). Moreover, LFCS10 achieved a 50.6% scan time reduction compared to SFGRAPPA and eliminated all aliasing artifacts observed in standard FOV imaging.

Notably, the proportion of scans with IQ scores ≥3 was higher for LFCS7 (35 of 40) and LFCS10 (36 of 40) compared to SFGRAPPA (33 of 40). This improvement may be attributed to the regularization inherent in CS, which suppresses noise and preserves edge detail, thereby enhancing perceived IQ. These findings further confirm that increasing the acceleration rate in LFCS10 does not compromise perceived image quality.

Our qualitative analyses showed a discordance between perceived SNR and IQ scores when comparing SFGRAPPA to LFCS7 or LFCS10. Specifically, the perceived SNR scores were significantly lower compared to SFGRAPPA’s SNR scores. However, the perceived IQ scores were not significantly different between SFGRAPPA vs LFCS7 or LFCS10. Other studies have similarly shown SNR to be a poor predictor of perceived diagnostic value (16). One possible explanation for the lower SNR scores could be due to the use of a 20-channel instead of a 64-channel head coil. With a higher number of channels, the 64-channel head coil provides a gain in SNR compared to the 20-channel head coil when using high acceleration factors. With the 64-channel head coil, however, the gain may be greatest at the surface with reduced sensitivity at further distances from the coil. This architecture can introduce signal-intensity inhomogeneity in the image and compromise uniformity in deeper brain regions (17, 18). The anatomy of interest for intracranial VWI is the circle of Willis, which sits centrally at the base of the brain. Thus, the gain in SNR using a 64-channel head coil may be incremental for VWI. Another study comparing 20-versus 64-channel head coils with and without receiving bias (B1-) normalization (e.g., prescan normalization filter) showed that for structural imaging, the 64-channel head coil with prescan normalization filter indeed provided higher SNR and CNR results than 20-channels (19). However, the prescan normalization filter had a stronger influence than the coil itself, as there was no significant difference in SNR or CNR between head coils without the prescan normalization filter (19). These results suggest normalization of signal-intensity inhomogeneity to produce clinically uniform images may have a greater impact to an radiologist.

At our institution, the 20-channel head coil is readily available across our inpatient and outpatient imaging centers and is larger in size (315 (length) x 475 (width) x 360 (height) mm) (20) than the 64-channel head coil (435 x 395 x 350 mm) (21). The 20-channel head coil’s larger size may result in higher patient tolerance, particularly in claustrophobic patients and is hence preferred in the clinical setting. Additionally, the goal of this study was to consider the range of small-to-large head sizes across our patient population to reduce potential parameter changes introduced by the technologist. Thus, we used a 20-channel head coil in this study. A future direction is to compare the 20-versus 64-channel head coils in IQ and SNR for the different acceleration techniques and evaluate the potential gains for intracranial VWI.

The primary goal of this study was to reduce scan times. We achieved an up to 50.6% reduction in scan time with CS compared to SFGRAPPA while maintaining 0.533mm spatial resolution, IQ and scanning a larger FOV. The larger FOV also allowed us to eliminate aliasing artifacts. A prior study using CS-SPACE with acceleration factors of 3x to 7x also achieved scan time reductions between ∼37% and ∼58% with whole-brain coverage in under 7 minutes while maintaining high IQ (5). Typically, IQ remained robust up to a 5x acceleration factor, but higher accelerations led to some image degradation (5). Zhu et al. attempted higher accelerations to reduce the scan time to around 4:31 minutes, but the reconstructions produced inaccurate wall area estimations (5). We explored CS with a 10x acceleration factor and achieved IQ comparable to the SOC SFGRAPPA with a scan time of 4 minutes 55 seconds. Despite compromises in SNR, we were still able to effectively visualize the vessel walls and lumen for LFCS10 with a 50.6% reduction in scan time compared to SFGRAPPA. However, it is important to note that these findings may be influenced by the relatively small sample size, and further validation with a larger cohort is warranted.

Our vessel segment analysis underscores that the BA is the most challenging vessel for consistent visualization of wall margins across all sequences. The deep location in the brain and distance from the head coil likely contribute to the signal degradation. Furthermore, there is high cerebrospinal fluid (CSF) flow variability around the basilar artery where CSF velocities can vary with cardiac pulsations (22, 23). The need for effective CSF flow suppression in the prepontine cistern due to pulsatility driven by the cardiac cycle can affect the conspicuity of the BA vessel wall. Techniques like anti-driven-equilibrium (28), delay alternating with nutation for tailored excitation (DANTE), and improved motion sensitized driven-equilibrium (iMSDE) have been investigated for optimal CSF and blood flow suppression and need to be systematically tested with different acceleration techniques (24, 25,27).

There are several limitations to this study. The study was conducted on a small sample of healthy volunteers with pre-contrast imaging only. Image comparisons in patients with intracranial vasculopathies using post-contrast VWI would be advantageous, given contrast enhancement of the vessel walls may further enhance conspicuity of pathologic vessel walls at reduced scan times. To minimize patient burden, we compared CS and CAIPI acceleration techniques in healthy volunteers to first identify the optimal acceleration technique before testing a combined post-gadolinium SOC and experimental protocol on patients. This reduces patient burden with a single scan exam. Second, we did not perform a head-to-head comparison of a large FOV CS vs GRAPPA accelerated sequence, given a large FOV GRAPPA-accelerated sequence would exceed 10 minutes. In addition to the lengthy acquisition time, images may be confounded by motion artifacts for scans longer than 10 minutes.

## Conclusion

This study aimed to optimize and accelerate the 3D T1-weighted VF-TSE sequence to accommodate varying head sizes, minimize aliasing artifacts, and achieve IQ comparable to a SOC GRAPPA accelerated sequence. The large FOV CS-accelerated sequence with 10x acceleration did not show any statistical difference in IQ compared to the standard FOV GRAPPA sequence. Despite a lower perceived SNR, all aliasing artifacts were eliminated. The large FOV CS-accelerated sequence at 10x acceleration thus offers clinical advantages with a reduced scan time at 4 minutes 55 seconds (a 50.6% reduction in scan time compared to SOC GRAPPA), eliminates aliasing artifact and accommodates varying head sizes.

## Data Availability

The data that support the findings of this study are available from the corresponding author upon reasonable request.

## Notes

Grant Support: JWS is supported by American Heart Association Career Development Award (938082) and Institute of Translational Medicine and Therapeutics; ZF is supported by R01HL147355; CZ is supported by R01HL162743.

### Competing Interest Statement

Disclosures: Manuel Taso and Robert Sellers are employed by Siemens Medical Solutions, USA

### Funding Statement

Grant Support: Jae W Song is supported by American Heart Association Career Development Award (938082) and Institute of Translational Medicine and Therapeutics; Zhaoyang Fan is supported by R01HL147355; Chengcheng Zhu is supported by R01HL162743.

### Author Declarations

The Institutional Review Board of the University of Pennsylvania provided ethical approval for this study.

## References

1. Mandell D, Mossa-Basha M, Qiao Y, et al. Intracranial vessel wall MRI: principles and expert consensus recommendations of the American Society of Neuroradiology. American Journal of Neuroradiology 2017;38(2):218–229.

2. Qiu Z, Jia S, Su S, et al. Wave-CAIPI Highly Accelerated Whole-Brain Intracranial Vessel Wall Imaging. Proc Intl Soc Mag Reson Med; 2020. p. 2133.

3. Breuer F, Blaimer M, Griswold M, Jakob P. Controlled aliasing in parallel imaging results in higher acceleration (CAIPIRINHA). Magnetom Flash 2012;1:2–7.

4. Kharaji M, Canton G, Guo Y, Mosi MH, Zhou Z, Balu N, Mossa-Basha M. DANTE-CAIPI Accelerated Contrast-enhanced 3D T1: Deep-Learning-Based Image Quality Improvement for Vessel Wall MRI. American Journal of Neuroradiology 2025;46(1):49–56.

5. Zhu C, Tian B, Chen L, et al. Accelerated whole brain intracranial vessel wall imaging using black blood fast spin echo with compressed sensing (CS-SPACE). Magnetic Resonance Materials in Physics, Biology and Medicine 2018;31:457–467.

6. Donoho DL. Compressed sensing. IEEE Transactions on information theory 2006;52(4):1289–1306.

7. Lustig M, Donoho DL, Santos JM, Pauly JM. Compressed sensing MRI. IEEE signal processing magazine 2008;25(2):72–82.

8. Fan Z, Yang Q, Deng Z, et al. Whole brain intracranial vessel wall imaging at 3 T esla using cerebrospinal fluid–attenuated T1 weighted 3 D turbo spin echo. Magnetic resonance in medicine 2017;77(3):1142–1150.

9. Beals KL, Smith CL, Dodd SM, et al. Brain size, cranial morphology, climate, and time machines [and comments and reply]. Current Anthropology 1984;25(3):301–330.

10. Ho K-C, Roessmann U, Straumfjord J, Monroe G. Analysis of brain weight. I. Adult brain weight in relation to sex, race, and age. Archives of pathology & laboratory medicine 1980;104(12):635–639.

11. Harvey I, Persaud R, Ron MA, Baker G, Murray R. Volumetric MRI measurements in bipolars compared with schizophrenics and healthy controls. Psychological Medicine 1994;24(3):689–699.

12. Technologists R. Radiologic Technologist Best Practices for MR Safety. American Society of Radiologic Technologists; 2018.

13. Qiao Y, Anwar Z, Intrapiromkul J, et al. Patterns and implications of intracranial arterial remodeling in stroke patients. Stroke 2016;47(2):434–440.

14. Pan Z, Qin Y, Bai W, He Q, Yin X, He J. Implementing multiple imputations for addressing missing data in multireader multicase design studies. BMC Medical Research Methodology 2024;24(1):217.

15. Fedorov A, Beichel R, Kalpathy-Cramer J, et al. 3D Slicer as an image computing platform for the Quantitative Imaging Network. Magnetic resonance imaging 2012;30(9):1323–1341.

16. Chapman BE, Goodrich CK, Alexander AL, Blatter DD, Parker DL. Evaluation of measures of technical image quality for intracranial magnetic resonance angiography. Computers and biomedical research 1999;32(6):530–556.

17. Lin FH, Chen YJ, Belliveau JW, Wald LL. A wavelet based approximation of surface coil sensitivity profiles for correction of image intensity inhomogeneity and parallel imaging reconstruction. Human brain mapping 2003;19(2):96–111.

18. Wiggins GC, Triantafyllou C, Potthast A, Reykowski A, Nittka M, Wald L. 32 channel 3 Tesla receive only phased array head coil with soccer ball element geometry. Magnetic Resonance in Medicine: An Official Journal of the International Society for Magnetic Resonance in Medicine 2006;56(1):216–223.

19. Schmitt T, Rieger JW. Recommendations of choice of head coil and prescan normalize filter depend on region of interest and task. Frontiers in neuroscience 2021;15:735290.

20. Healthineers S. Tx/Rx CP Head Coil. In: Healthineers S, editor. Magnetic Resonance Imaging-Coils. https://www.siemens-healthineers.com/en-us/magnetic-resonance-imaging/options-and-upgrades/coils/tx-rx-cp-head-coil; 2024.

21. Healthineers S. Head/Neck 64. In: Healthineers S, editor. Magnetic Resonance Imaging-Coils: https://www.siemens-healthineers.com/en-us/magnetic-resonance-imaging/options-and-upgrades/coils/64-channel-head-neck-coil; 2024.

22. Wagshul ME, Chen JJ, Egnor MR, McCormack EJ, Roche PE. Amplitude and phase of cerebrospinal fluid pulsations: experimental studies and review of the literature. Journal of neurosurgery 2006;104(5):810–819.

23. Yamada S, Tsuchiya K, Bradley W, et al. Current and emerging MR imaging techniques for the diagnosis and management of CSF flow disorders: a review of phase-contrast and time–spatial labeling inversion pulse. American journal of neuroradiology 2015;36(4):623–630.

24. Sannananja B, Zhu C, Colip C, et al. Image-quality assessment of 3D intracranial vessel wall MRI using DANTE or DANTE-CAIPI for blood suppression and imaging acceleration. American Journal of Neuroradiology 2022;43(6):837–843.

25. Kim D, Lee H-J, Baik J, Hwang M, Miyoshi M, Kang Y. Improved blood suppression of motion-sensitized driven equilibrium in high-resolution whole-brain vessel wall imaging: comparison of contrast-enhanced 3D T1-weighted FSE with motion-sensitized driven equilibrium and delay alternating with nutation for tailored excitation. American Journal of Neuroradiology 2022;43(12):1713–1718.

26. Yang Q et al. Whole-Brain Vessel Wall MRI: A ParameterTune-up Solution to Improve the ScanEfficiency of Three-Dimensional VariableFlip-Angle Turbo Spin-Echo. J. MAGN. RESON. IMAGING 2017;46:751–757

27. Kong Q, Xiao J, Shiroishi MS, Sheikh-Bahaei N, Cen SY, Khatibi K, Mack WJ, Ye JC, Kim PE, Bi X, Saloner D, Yang Q, Chang E, Fan Z. Interleaved flow-sensitive dephasing (iFSD): Toward enhanced blood flow suppression and preserved T_1_ weighting and overall signals in 3D TSE-based neuroimaging. Magn Reson Med. 2025 May;93(5):1911–1923. doi: 10.1002/mrm.30391. Epub 2024 Dec 8. PMID: 39648519; PMCID: PMC11893033.

28. Yang H, Zhang X, Qin Q, Liu L, Wasserman BA, Qiao Y. Improved cerebrospinal fluid suppression for intracranial vessel wall MRI. J Magn Reson Imaging. 2016 Sep;44(3):665–72. doi: 10.1002/jmri.25211. Epub 2016 Mar 7. PMID: 26950926.

