## Supplemental Table 1 and 2 for "Optimizing scan efficiency of T1-weighted imaging for whole-brain intracranial vessel wall imaging"

Supplemental Table 1. VWI Examples for Scoring

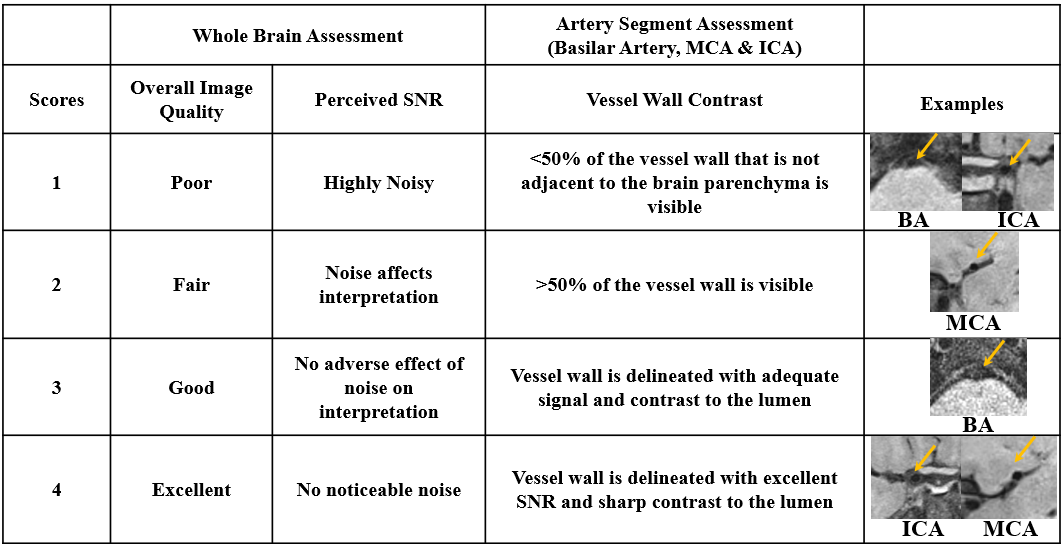

Abbreviations: BA, basilar artery; MCA, middle cerebral artery; ICA, internal carotid artery

Supplemental Table 2: Quantitative Measurements of the Actual Measured and Perceived Lumen and Wall Diameters

| **Vessel Wall/Lumen Segment** | **SFGRAPPA**  **(SOC)** | **SFCAIPI** | **SFCS7** | **LFCS7** | **LFCS10** |
| --- | --- | --- | --- | --- | --- |
| BA- Lumen | 2.89 ± 0.55 | 2.72 ± 0.41 | 3.19 ± 0.35* | 2.99 ± 0.27 | 2.97 ± 0.25 |
| BA- Wall | 4.77 ± 0.40 | 4.24 ± 0.68* | 4.92 ± 0.54 | 4.62 ± 0.44 | 4.90 ± 0.36 |
| MCA- Lumen | 2.58 ± 0.35 | 2.76 ± 0.47 | 2.57 ± 0.18 | 2.85 ± 0.34* | 2.60 ± 0.43 |
| MCA- Wall | 4.03 ± 0.49 | 4.24 ± 0.35 | 4.06 ± 0.34 | 4.35 ± 0.45* | 4.02 ± 0.61 |
| ICA- Lumen | 3.44 ± 0.79 | 3.54 ± 0.44 | 3.71 ± 0.40 | 3.60 ± 0.58 | 3.51 ± 0.66 |
| ICA- Wall | 5.29 ± 0.90 | 5.22 ± 0.54 | 5.49 ± 0.57 | 5.64 ± 0.58 | 5.36 ± 0.71 |

*p<0.05

Abbreviations: SD, standard deviation; SOC, standard of care; SFGRAPPA, standard FOV GRAPPA R=2; SFCAIPI, standard FOV CAIPI R=4; SFCS7, standard FOV CS R=7; LFCS7, large FOV CS7; LFCS10, large FOV CS10 R=10; BA, basilar artery; MCA, middle cerebral artery; ICA, internal carotid artery
